# Breastfeeding and related feeding practices during the first 6 months of life in seven countries across Asia and Africa: a cross-sectional survey

**DOI:** 10.1101/2025.11.18.25340526

**Authors:** Ameena Goga, BRANCH pre-intervention infant feeding survey study group

## Abstract

Despite investments in breastfeeding, and evidence illustrating that breastfeeding has short- and long-term health and economic benefits, countries are far off from global breastfeeding targets. To inform a breastfeeding counselling and support intervention within a randomized community-based trial titled Breastfeeding Counselling and Management of Growth Faltering in Infants Aged under Six Months (BRANCH in seven low-middle income countries: Bangladesh, India, Pakistan, Ethiopia, Nigeria, Tanzania, and Uganda we conducted a cross-sectional home-based infant feeding survey in each country (September-December 2023).

We enrolled 4,308 infants aged 0-6 months. Among 1378 infants aged 60 days or younger, almost all (97.3%, 95% CI 96.3-98.1%) received mothers own milk; however, early breastfeeding initiation within the first hour of birth (EIBF) occurred in only half (50.8%, 95% CI 48.1-53.6%), with marked regional differences – two-thirds in Africa (66.5% (95% CI 62.9-69.9%) versus one-third in Asia (31.8% (95% CI 28.1-35.7%). EBF prevalence was only 52.9% (95% CI 51.4-54.4%) and 40.2% (95% CI 38.7-41.7%), by 24 hour and 7-days recall, respectively. The foods and fluids that interrupted EBF varied by country. Since birth almost one-quarter of infants (23.9%, 95% CI 22.7-25.2%) had ever received formula milk, with higher use in Asia (31.6%, 95% CI 29.5-33.8%) than Africa (18.4%, 16.9-20.0. Most formula use started within the first month of life (65.0%), predominantly due to perceived insufficient breast milk (64.7%) and advice from healthcare providers (52.0%). Strikingly, only 46.9% of mothers using formula milk prepared it correctly, with substantial variation across sites (8.3% in Bangladesh versus 77.0% in Ethiopia).

There is a critical need for context-specific breastfeeding counselling and support strategies for early and exclusive breastfeeding. These should build mothers’ and families’ confidence, strengthen breastfeeding support within the heath system and curb unnecessary formula milk marketing that exploits the fears, and concerns of parents and families.

## INTRODUCTION

In 2023, an estimated 4.8 million children died before the age of five, mostly from infectious diseases, undernutrition, and poor growth.(1) Most of these deaths are preventable with simple, cost-effective interventions. Breastfeeding is one of the most effective intervention to prevent under five mortality.(2, 3) Breast milk is optimal, complete food for infants, offering essential nutrition and robust immunological protection (4, 5) Breastfeeding provides multiple well-documented benefits for infant survival and development: It acts as an infant’s first food and vaccine, prevents both undernutrition and obesity, (2), prevents infectious diseases, and long-term non-communicable diseases and supports cognitive development.(6–9).

Breastfed newborns and infants aged 0-6 months have a four and 14-fold higher chance of survival, respectively, compared with non-breastfed newborns and 0-6 month infants, (10) and significantly reduces respiratory, diarrheal, and neonatal infections.(11) . Beyond its health benefits, breastfeeding is highly cost-effective, generating approximately $35 in return for every dollar invested, while inadequate investment in breastfeeding may cost the global economy approximately $507 billion annually.(12)

Consequently, WHO and United Nations Children’s Fund (UNICEF) recommend early breastfeeding initiation (EIBF- within the first hour of birth) and exclusive breastfeeding (EBF) for infants aged 0-6 months. (13) EBF is defined as ‘the infant receives only breast milk (including milk expressed or from a wet nurse) and no other liquids or solids, not even water, except for oral rehydration solution (ORS), drops or syrups of vitamins, minerals, or medicines prescribed by a health worker’. (14) Compared with predominant or partial breastfeeding, EBF provides superior nutrition and health benefits, strengthens the maternal-infant bond, and supports healthy birth spacing (6,7). WHO recommends at least six high quality breastfeeding counselling sessions during pregnancy and postpartum to promote EBF. (15)

However, global progress remains modest. EBF prevalence increased from 37% in 2012 to 48% in 2022 – an annual increase of only approximately 1%, (2), and only 46% of newborns are breastfed within the first hour of life.(13) Compared with other regions, EBF prevalence is lowest in Sub-Saharan Africa (36%) and South Asia (39%).(13) Disaggregated analysis of Demographic and Health Survey (DHS) data from five South Asian countries reveals EBF prevalence of 58.5% in Bangladesh (2017–2018), 58.5% in India (2019–2021) and 42.4% in Pakistan (2017–2018).(16) Targeted, tailored additional investments are needed to accelerate progress and achieve the World Health Assembly (WHA) target of at least 50% EBF in infants aged 0-6 months by 2025 (17), and the Global Breastfeeding Collective’s target of ≥70% 0-6 month EBF prevalence by 2030. (13)

In South Asia and Sub-Saharan Africa, poor breastfeeding practices stem from a mix of socio-cultural, economic, educational, and health system barriers.(18) Cultural practices such as discarding colostrum and early introduction of other liquids or solids often delay breastfeeding initiation.(19) Additional challenges include limited awareness of the critical role of breastfeeding for child health, short maternity leave, lack of community support and maternal malnutrition.(7, 11) Economic pressures, especially for women in informal jobs, and weak health systems further disrupt breastfeeding.(20) The situation is further exacerbated by the uncontrolled marketing of breastmilk substitutes, in violation of the International Code of Marketing of Breastmilk Substitutes and this fosters misconceptions about the safety of formula feeding.(21)

Cognizant of the importance of breastfeeding, a randomized community-based trial titled Breastfeeding Counselling and Management of Growth Faltering in Infants Aged under Six Months (BRANCH) is underway in seven low-middle income countries (LMICs): Bangladesh, India, Pakistan, Ethiopia, Nigeria, Tanzania, and Uganda ((ANZ Clinical trial registry, CTRN12624000704594). BRANCH aims to determine the effect of nutritional supplementation plus intensified breastfeeding counselling and support (IBFCS) delivered antenatally and postnatally at household level, compared with IBFCS alone on mortality, morbidity and growth in infants aged 0-6 months with growth faltering. Here, we report findings from a baseline survey conducted to understand breastfeeding and related practices, including early initiation, exclusivity (EBF), use of prelacteal feeds, formula / other milk consumption, and the use of solid / soft foods at BRANCH trial sites.

## METHODS

### Methods

#### Study Design and Setting

A cross sectional, community-based age-stratified survey was conducted, among randomly selected mothers of young infants aged 0-6 months living in the BRANCH trial catchment area. Thus the study sites were located in Bangladesh - rural Zakiganj sub-district of Sylhet district, northeast region; India - urban and peri-urban areas in South Delhi, including Sangam Vihar, Tigri, Madangir, Dakshinpuri and Khanpur; Pakistan - peri-urban, coastal Karachi (Ibrahim Hyderi and Ali Akber Shah Goth), Sindh; Ethiopia - urban districts of Addis Ababa (Yeka), Nigeria - urban areas within Ife local government, south west region, Osun state, Tanzania - all urban and rural districts of Pemba island and Uganda - rural Iganga and Mayuge districts, in the eastern region.

#### Study population

All mothers with young infants aged 0-6 months residing in the catchment area were eligible for enrolment. Twins were included but other multiple births, infants with a cleft palate or any other birth condition that may affect feeding, such as congenital heart disease were excluded.

#### Sample size and sampling technique

Based on the assumption that 50% of the population would be exclusively breastfeeding in the first six months, a sample size of 600 infants per country would enable the detection of EBF point prevalence with an accuracy of ±4%. Assuming 85% initiation of breastfeeding within one hour at most sites, a sample size of 200 infants aged less than two months would enable detection of point prevalence with an accuracy of +/- 5%. A priori, it was decided to recruit into six age groups: 0-30 days, 31-60 days, 61-90 days, 91-120 days, 121-150 days, and 151–180 days, with 100 infants per age group per site. Simple random sampling was used. After determining the sample size, random selection of an enlisted household was achieved using a random number generator. These households were then approached for participation in the survey.

#### Data Collection

Data collection occurred from 18th September-31^st^ December 2023). The questionnaire was developed after reviewing data collection tools from Demographic and Health Surveys (DHS), Multiple Indicator Cluster Surveys, (MICS) and WHO survey tools. Trained field assistants visited each household, explained the survey, enquired about women with infants less than 6 months of age, and offered them participation. In cases where the mother and/or infant were temporarily away (house was locked, mother / infant still hospitalized after delivery), two revisits were made within 5 and 10 days of the first visit. Families who had relocated out of the BRANCH area, and mothers not available on the 3^rd^ visit were screened out.

Following individual, written informed consent in local languages, involving impartial witnesses and thumbprints for illiterate participants, trained field assistants administered the survey questionnaire to mothers / primary caregivers in local languages. Details about breastfeeding, use (including preparation) of formula milk, use of other milk and foods, reasons for introducing formula / other milks, and method of feeding were collected for all enrolled infants. Data on infant feeding practices were based on recall over three-time intervals: in the last 24-hours, in the last 7-days preceding the interview and since birth of the infant (‘ever’ questions). Data on immediate feeding after birth (early initiation of breastfeeding (EIBF), prelacteals) were restricted to mothers of infants < 120 days of age to limit recall bias. Infant height and weight, and mid-upper arm circumference (MUAC) weremeasured following standardized training, using Seca 354, Seca 457 and Seca 212. Infants who were sick or with WLZ < −2SD or were referred to the nearest health facility.

Field assistants were trained, using standardised operating procedures (SOPs), on study conduct and how to administer the questionnaire, to ensure consistency across all seven sites. Monthly site meetings were conducted to support consistency.

Data were collected either electronically on tablets or using paper-case report forms (CRFs). Sites using paper CRFs entered data electronically within 7-days. Data were cleaned and, range and logical checks applied. All changes were stored in the audit trail.

All data were anonymized by removing identifying participant (including household) or site information. WHO received cleaned anonymised data from each site.

##### Ethics

Ethics approval was obtained from the WHO Ethics Review Committee (ERC.0003929), and from local ethics committees overseeing all the seven sites (Projahnmo Research Foundation Institutional Review Board (Bangladesh), Addis Continental Institute of Public Health Institutional Research Ethics Committee and Harvard T.H. Chan School of Public Health (Ethiopia), Society for Applied Studies (India), Obafemi Awolowo University Teaching Hospital Complex Ethics Research Committee (Nigeria), the Aga Khan University Ethics Review Committee (Pakistan), Zanzibar Health Research Institute (Pemba site, Zanzibar) and Makerere University College of Health Sciences School of Medicine Research Ethics Committee (Uganda). Identifiable data are retained securely, under lock and key, until the files are clean, analysis completed, and results published. Deidentified data will be stored permanently. The cleaned anonymized research dataset was sent to WHO by all the sites.

### Data Analysis

Data collected at each of the sites were shared with, and combined by, the BRANCH survey analysis team. Range and consistency checks were run on individual sites’ datasets, and consistency checks were run across the datasets to ensure harmonised coding of variables. Inconsistencies were checked and cleaned in collaboration with individual sites. The analyses were conducted using Stata 18.1 (StataCorp. 2023. Stata Statistical Software: Release 18. College Station, TX: StataCorp LLC). Percentages for categorical variables, reporting point prevalence, with 95% confidence intervals [95% CI] were estimated using the exact Clopper-Pearson method, and means with standard deviations for continuous variables, were calculated for each country site, combined for the Asian and African sites (separately), and combined overall. Data completeness was high overall (<1% missing data for most variables). Outcomes were defined, apriori, as in Text Box 1.

##### Text Box 1: Outcomes variables

**Early initiation of breastfeeding (EIBF):** Provision of breastmilk to the infant within one hour of birth.

**EBF:** the infant receives only breast milk (including milk expressed or from a wet nurse) and no other liquids or solids, not even water, except for oral rehydration solution (ORS), drops or syrups of vitamins, minerals, or medicines prescribed by a health worker

**Early introduction of complementary food:** Introduction of any food (liquid/ solid) other than breastmilk before the completion of 6 months (180 days).

**Formula milk use:** Any reported use of formula milk to feed the infant. In this study, operationalised as proportion of infants who were given formula milk in the last 24 hrs or 7-days, depending on which question is being reported on, out of the total number of infants surveyed. For rates across each strata proportion of infants given formula milk in each strata out of the total number of infants in that strata.

**Animal milk use:** Any reported use of animal milk to feed the infant. In this study, operationalised as proportion of infants who were given animal milk the last 24 hrs or 7-days, depending in which question is being reported on, out of the total number of infants surveyed. For rates across each strata proportion of infants given animal milk in each strata out of the total number of infants in that strata.

**Use of prelacteals**: infant aged less than 2 months and received any other foods, fluids or medicines in the first 72 hours of birth .

The correct preparation of formula milk was self-reported, in response to four questions: whether the mother always (i) washes her hands (ii) boils the water (iii) sterilises the utensils before preparing a feed, and (iv) whether she always prepares one need at a time. A yes response to all these questions contributed to a yes response to ‘correct preparation of formula milk’.

## Results

We recruited 4,308 mother-infant pairs, 42% from Asia and 58% from Africa (Table 1); mothers were on average 27.5 (standard deviation (SD) ±5.7) years old with 9 years (±4.3) of schooling; working at home (homemaker / housewife) was far commoner in Asia (92%) compared with Africa (43%), Table 1. Almost all mothers (95.4%) reported previous breastfeeding experience, and antenatal care (96.8%) during this pregnancy; infants were mostly singleton and facility-born, with low birth weight and preterm delivery commoner in Asia (30% and 21% respectively), compared with Africa (15% and 3% respectively); wasting prevalence was similar across regions but stunting and underweight for age were almost twice as high in Asia compared with Africa, Table 1.

**Table 1:** Baseline characteristics of mother-infant pairs.

|  | All sites | Asia |  |  |  | Africa |  |  |  |  |
| --- | --- | --- | --- | --- | --- | --- | --- | --- | --- | --- |
|  |  | Bangladesh | India | Pakistan | Total | Ethiopia | Nigeria | Tanzania | Uganda | Total |
| **Total** | **4308** | 600 | 600 | 600 | **1800** | 625 | 649 | 600 | 634 | **2508** |
| **Maternal/ respondent's characteristics** |  |  |  |  |  |  |  |  |  |  |
| Mean Age in years (SD) | **27.5 (5.7)** | 25.6 (5.0) | 27.4 (4.4) | 26.6 (5.6) | **26.5 (5.1)** | 29.3 (4.8) | 28.6 (6.2) | 29.2 (6.1) | 25.9 (6.1) | **28.3 (6.0)** |
| Mean years of schooling* (SD) | **9.0 (4.3)** | 8.7 (3.1) | 10.6 (4.6) | 5.0 (4.3) | **8.1 (4.7)** | 10.1 (4.4) | 11.7 (3.0) | 8.8 (3.5) | 7.8 (3.2) | **9.6 (3.9)** |
| Housewife/home maker, N (%) | **2735 (63.5)** | 593 (98.8) | 570 (95.0) | 497 (82.8) | **1660 (92.2)** | 417 (66.7) | 0 | 402 (67.0) | 256 (40.5)<sup>ys</sup> | **1075 (42.9)** |
| **Of those who work, what is their occupation?** | **N=1571** | **N=7** | **N=30** | **N=103** | **N=140** | **N=208** | **N=649** | **N=198** | **N=376** | **N=1431** |
| Government job | **701 (44.6)** | 3 (42.9) | 3 (10.0) | 2 (1.9) | 8 (5.7) | 56 (26.9) | 604 (93.1) | 26 (13.1) | 7 (1.9) | 693 (48.4) |
| Private job | **201 (12.8)** | 3 (42.9) | 15 (50.0) | 2 (1.9) | 20 (14.3) | 99 (47.6) | 37 (5.7) | 16 (8.1) | 29 (7.7) | 181 (12.7) |
| Self employed | **402 (25.6)** | 0 | 12 (40.0) | 52 (50.5) | 64 (45.7) | 46 (22.1) | 8 (1.2) | 101 (51.0) | 183 (48.7) | 338 (23.6) |
| Farming | **198 (12.6)** | 0 | 0 | 1 (1.0) | 1 (0.7) | 0 | 0 | 49 (24.8) | 148 (39.4) | 197 (13.8) |
| Daily wage laborer | **57 (3.6)** | 1 (14.3) | 0 | 46 (44.7) | 47 (33.6) | 5 (2.4) | 0 | 1 (0.5) | 4 (1.1) | 10 (0.7) |
| Other | **12 (0.8)** | 0 | 0 | 0 | 0 | 2 (1.0) | 0 | 5 (2.5) | 5 (1.3) | 12 (0.8) |
| House has electricity, N (%) | **3982 (92.4)** | 599 (99.8) | 600 (100) | 597 (99.5) | **1796 (99.8)** | 621 (99.4) | 628 (96.8) | 408 (68.0) | 592 (83.4) | **2186 (87.2)** |
| Main water source is an improved water source, N (%) | **4179 (97.0)** | 495 (82.5) | 599 (99.8) | 598 (99.7) | **1692 (94.0)** | 625 (100) | 647 (99.7) | 588 (98.0) | 627 (98.9) | **2487 (99.2)** |
| Improved sanitation, N (%) | **4211 (97.8)** | 594 (99.0) | 600 (100) | 599 (99.8) | **1793 (99.6)** | 587 (93.9) | 627 (96.6) | 574 (95.7) | 630 (99.4) | **2418 (96.4)** |
| Clean fuel for cooking, N(%) | **2314 (53.7)** | 33 (5.5) | 599 (99.8) | 577 (96.2) | **1209 (67.2)** | 492 (78.7) | 601 (92.6) | 10 (1.7) | 2 (0.3) | **1105 (44.1)** |
| **Past pregnancy and breastfeeding history of mother** |  |  |  |  |  |  |  |  |  |  |
| Mean number of liveborn Infants (SD) | **2.8 (1.9)** | 2.3 (1.2) | 1.9 (1.0) | 3.2 (2.2) | **2.5 (1.7)** | 2.1 (1.1) | 2.4 (1.3) | 4.4 (2.6) | 2.9 (2.0) | **2.9 (2.0)** |
| Previous BF experience, N (%)<sup>φ</sup> | **2842 (95.4)** | 393 (99.5) | 354 (96.2) | 433 (98.9) | **1180 (98.3)** | 407 (96.2) | 336 (80.4) | 516 (100) | 403 (96.0) | **1662 (93.5)** |
| Mother attended antenatal care N (%)<sup>†</sup> | **4167 (96.8)** | 530 (88.3) | 591 (98.7) | 583 (97.2) | **1704 (94.7)** | 621 (99.4) | 619 (95.4) | 598 (99.7) | 625 (98.7) | **2463 (98.2)** |
| **Infant characteristics** |  |  |  |  |  |  |  |  |  |  |
| Female sex at birth | **2116 (49.1)** | 298 (49.7) | 309 (51.5) | 283 (47.2) | **890 (49.4)** | 290 (46.4) | 329 (50.7) | 288 (94.0) | 319 (50.3) | **1226 (48.9)** |
| Singleton birth | **4199 (97.5)** | 578 (96.3) | 582 (97.0) | 590 (98.3) | **1750 (97.2)** | 605 (96.8) | 626 (96.5) | 592 (98.7) | 626 (98.7) | **2449 (97.7)** |
| **Place of delivery** |  |  |  |  |  |  |  |  |  |  |
| Health facility | **3530 (82.0)** | 337 (56.2) | 565 (94.2) | 467 (77.8) | **1369 (76.1)** | 620 (99.2) | 460 (71.0) | 544 (90.7) | 537 (84.7) | **2161 (86.2)** |
| Home | **598 (13.9)** | 262 (43.7) | 33 (5.5) | 132 (22.0) | **427 (23.7)** | 5 (0.8) | 73 (11.3) | 48 (8.0) | 45 (7.1) | **171 (6.8)** |
| Other | **179 (4.1)** | 1 (0.2) | 2 (0.3) | 1 (0.2) | **4 (0.2)** | 0 | 115 (17.8) | 8 (1.3) | 52 (8.2) | **175 (7.0)** |
| Not known | **1** | 0 | 0 | 0 | **0** | 0 | 1 | 0 | 0 | **1** |
| Mean infant age in days at the time of survey administration (SD) | **94.5 (51.9)** | 90.8 (51.3) | 98.6 (52.5) | 97.0 (51.7) | **95.5 (51.9)** | 105.5 (47.2) | 90.1 (52.9) | 89.7 (52.9) | 90.1 (52.6) | **93.8 (51.9)** |
| Mean birth weight of infant (SD)<sup>†</sup> | **3.0 (0.6)** | 2.9 (0.5) | 2.8 (0.5) | 2.8 (0.6) | **2.8 (0.6)** | 3.2 (0.5) | 3.2 (0.7) | 3.2 (0.6) | 3.2 (0.7) | **3.2 (0.6)** |
| Low birth weight, N (%)<sup>†</sup> | **738 (21.2)** | 140 (32.1) | 161 (27.6) | 129 (31.6) | **430 (30.1)** | 81 (13.9) | 67 (19.7) | 70 (12.0) | 90 (16.6) | **308 (15.0)** |
| Baby born at <9 months gestation, N (%)<sup>‡</sup> | **456 (10.6)** | 28 (4.6) | 209 (34.8) | 146 (24.3) | **383 (21.3)** | 17 (2.7) | 18 (2.8) | 8 (1.3) | 30 (4.8) | **73 (2.9)** |
| Underweight for age, N (%)<sup>§</sup> | **690 (16.3)** | 100 (16.7) | 118 (20.0) | 197 (33.9) | **415 (23.3)** | 44 (7.46) | 124 (19.1) | 70 (11.78) | 37 (5.9) | **275 (11.2)** |
| Stunted, N (%)<sup>§</sup> | **671 (15.8)** | 89 (14.8) | 102 (17.0) | 181 (31.1) | **372 (20.9)** | 38 (6.4) | 94 (14.5) | 113 (19.0) | 54 (8.6) | **299 (12.1)** |
| Wasted N (%)<sup>§</sup> | **463 (11.0)** | 62 (10.4) | 54 (9.1) | 74 (12.9) | **190 (10.7)** | 85 (14.6) | 134 (20.7) | 28 (4.7) | 26 (4.2) | **273 (11.2)** |

\* Defined as Z score below -5 SD or above +5 SD <sup>‡</sup>4 missing in Uganda <sup>§</sup>missing data n=39, 3 from Asian sites and 36 from African sites <sup>†</sup>missing data from India and Uganda n=1 each <sup>†</sup>missing data on birth weight, as follows: 372 in Asian sites (Bangladesh: 164; India:16; Pakistan 192) and 458 in African sites (Ethiopia: 42, Nigeria: 308; Tanzania:17; Uganda:91) <sup>†</sup>missing data on 5 babies: Ethiopia: 1; Uganda: 4 Underweight for age: 215 for age <-2 z score; Stunted: (Length for age score <-2 z-score); Wasted: (Weight for length z score <-2 z-score);, <sup>§</sup> missing or implausible values - Implausible defined as Z score below -6 SD or above +5 SD – for underweight: India: 3,Pakistan: 18, Ethiopia 35; Tanzania: 6; Uganda: 4; Stunted: India: 1; Pakistan: 18; Ethiopia: 30; Nigeria: 1; Tanzania: 5; Uganda: 6; wasted: Bangladesh:1; India: 4; Pakistan: 26; Ethiopia:44; Nigeria:3; Tanzania: 9; Uganda:16). <sup>§</sup>missing data on 2 participants

Among 1378 infants aged 60 days or younger, almost all (97.3%, 95% CI 96.3-98.1%) received mothers own milk, Table 2. However, early breastfeeding initiation within the first hour of birth (EIBF) occurred in only half (50.8%, 95% CI 48.1-53.6%), with marked regional differences – two-thirds in Africa (66.5% (95% CI 62.9-69.9%) versus one-third in Asia (31.8% (95% CI 28.1-35.7%); country-level variations were striking (Table 2): less than one-quarter of infants in India and Pakistan received EIBF (22.5% (95% CI 16.9-28.9%) and 25% (95% CI 19.4-31.9%), respectively), compared with over 80% in Tanzania and Uganda (82.8% (95% CI 76.6- 87.9%) and 88.8% (95% CI 83.6-92.8%), respectively), Table 2.

**Table 2:** Breastfeeding initiation in infants 60 days or younger.

|  | All sites | Asia |  |  |  | Africa |  |  |  |  |
| --- | --- | --- | --- | --- | --- | --- | --- | --- | --- | --- |
|  |  | Bangladesh | India | Pakistan | Total | Ethiopia | Nigeria | Tanzania | Uganda | Total |
| Number of infants aged 60 days or younger | 1378 | 200 | 200 | 200 | 600 | 131 | 224 | 204 | 219 | 778 |
| Number of infants who were given mothers' own breastmilk since birth, N (%) | 1341 (97.3%) | 200 (100%) | 200 (100%) | 198 (99.0%) | 598 (99.7%) | 127 (97.0%) | 216 (96.4%) | 188 (92.2%) | 212 (96.8%) | 743 (95.5%) |
| Breastfeeding initiation in <1 hour amongst BF infants, N (%) | 674 (50.8) | 95 (47.5) | 45 (22.5) | 50 (25.3) | 190 (31.8) | 64 (51.6) | 84 (39.4) | 154 (82.8) | 182 (88.8) | 484 (66.5) |
| Timing of breastfeeding Initiation after birth, N (%) |  |  |  |  |  |  |  |  |  |  |
| < 1 hour | 674 (50.8) | 95 (47.5) | 45 (22.5) | 50 (25.3) | 190 (31.8) | 64 (51.6) | 84 (39.4) | 154 (82.80) | 182 (88.8) | 484 (66.5) |
| 1 to <6 hours | 435 (32.8) | 90 (45.0) | 96 (48.0) | 76 (38.4) | 262 (43.8) | 42 (33.9) | 89 (41.8) | 23 (12.37) | 19 (9.3) | 173 (23.8) |
| 6 to <12 hours | 61 (4.6) | 7 (3.5) | 9 (4.5) | 23 (11.6) | 39 (6.5) | 1 (0.8) | 11 (5.2) | 9 (4.84) | 1 (0.5) | 22 (3.0) |
| 12 - <24-hours | 31 (2.3) | 4 (2.0) | 13 (6.5) | 8 (4.0) | 25 (4.2) | 4 (3.2) | 2 (0.9) | 0 | 0 | 6 (0.8) |
| 24+ hours | 125 (9.4) | 4 (2.0) | 37 (18.5) | 41 (20.7) | 82 (13.7) | 13 (10.5) | 27 (12.7) | 0 | 3 (1.5) | 43 (5.9) |
| Not known | 15 | 0 | 0 | 0 | 0 | 3 | 3 | 2 | 7 | 15 |

Almost all infants received some breastmilk in the last 24-hours (96.5%, 95% CI 96.0-97.1%) and 7-days (97.7%, 95% CI 97.2-98.1%), Table 3, Figures 1a/1b, Supplementary figures S1a-f. Yet EBF prevalence was only 52.9% (95% CI 51.4-54.4%) and 40.2% (95% CI 38.7-41.7%), by 24 hour and 7-days recall, respectively, Table 3, Figures 1a/1b, Supplementary figures S1a-f , with African sites reporting substantially higher prevalence (62.6% (95% CI 60.7-64.5%), and 47.9% (95% CI 45.9-49.8%, respectively) versus Asian sites (39.4% (95% CI 37.1-41.7%) and 29.4% (95% CI 27.4-31.6%), respectively). The foods and fluids that interrupted EBF varied by country, Table 3; in Bangladesh - solid/soft food, followed by water-based liquids and then formula milk; in India and Pakistan, other milk, followed by formula milk and solid/soft food, in Ethiopia, formula milk, in Nigeria, formula milk with an equal amount of water-based liquids and solid/soft food, then other milk, and in Tanzania and Uganda, almost equal use of water-based liquids and soft/solid food, with lower use of formula milk.

**Figure 1a:**
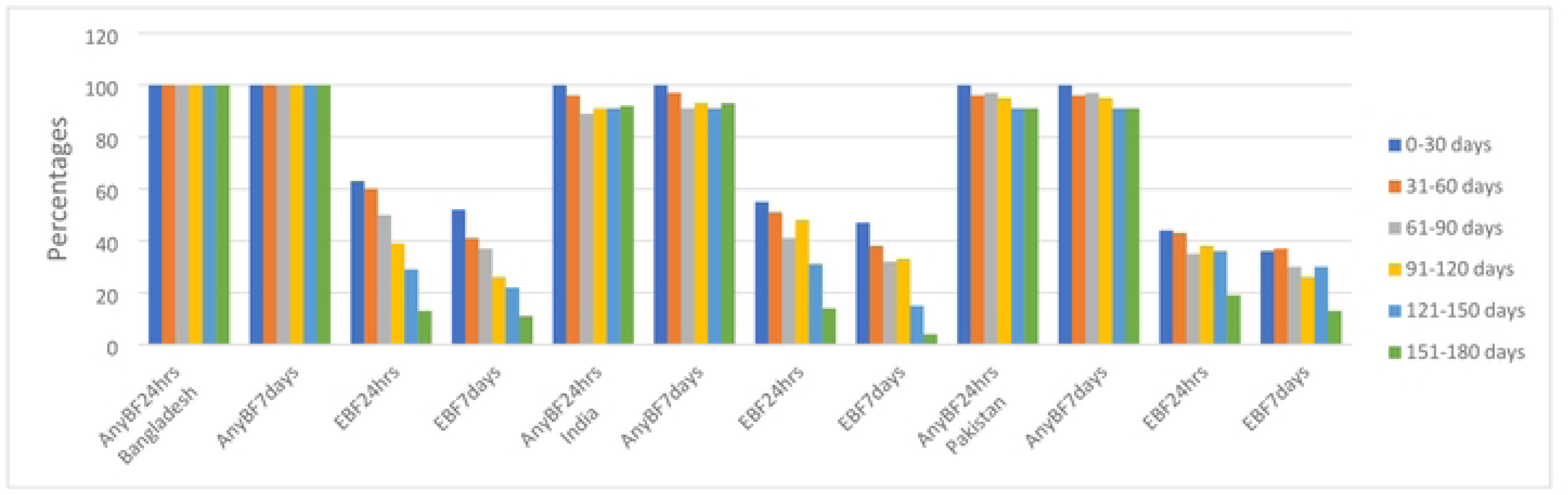
Percent practicing any breastfeeding or exclusive breastfeeding in the past 24-hours and 7-days by age group in the 3 Asian sites.

**Figure 1b:**
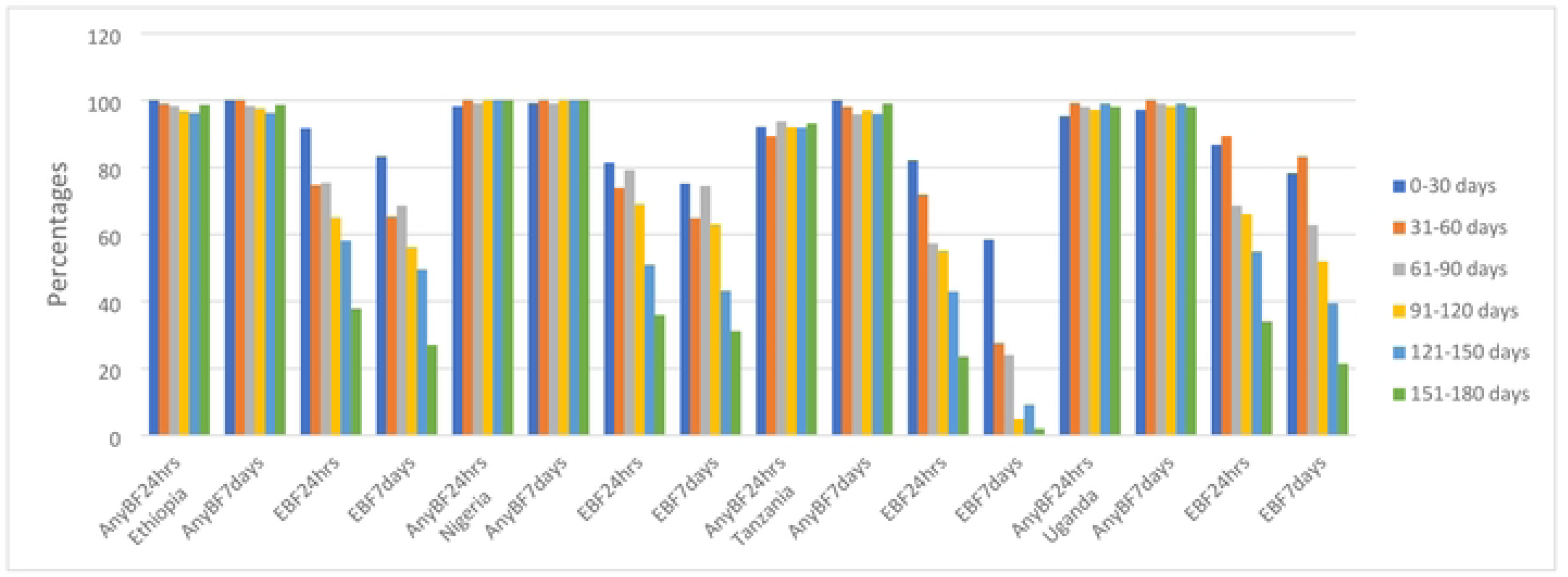
Percent practicing any breastfeeding or exclusive breastfeeding in the past 24-hours and 7-days by age group in the 4 African sites.

**Table 3:** Feeding in the last 24-hours and 7-days by age group, Number (%)

|  | All sites | Asia |  |  |  | Africa |  |  |  |  |
| --- | --- | --- | --- | --- | --- | --- | --- | --- | --- | --- |
|  |  | Bangladesh | India | Pakistan | Total - Asia | Ethiopia | Nigeria | Tanzania | Uganda | Total - Africa |
| **Total (all mother-infant pairs)** | **4308** | **600** | **600** | **600** | **1800** | **625** | **649** | **600** | **634** | **2508** |
| **Breastfeeding (BF) in last 24-hours** | **4159 (96.5)** | 600 (100) | 559 (93.2) | 570 (95.0) | **1729 (96.1)** | 612 (97.9) | 646 (99.5) | 552 (92.0) | 620 (97.8) | **2430 (96.9)** |
| **BF in last 7-days** | **4206 (97.7)** | 600 (100) | 565 (94.2) | 570 (95.0) | **1735 (96.4)** | 614 (98.2) | 647 (99.7) | 585 (97.7) | 625 (98.6) | **2471 (98.6)** |
| **Exclusive BF in last 24hrs** | **2280 (52.9)** | 254 (42.3) | 240 (40.0) | 215 (35.8) | **709 (39.4)** | 390 (62.4) | 423 (65.2) | 333 (55.5) | 425 (67.0) | **1571 (62.6)** |
| **Exclusive BF in last 7-days\*** | **1730 (40.2)** | 189 (31.5) | 169 (28.2) | 172 (28.7) | **530 (29.4)** | 334 (53.4) | 381 (58.7) | 126 (21.0) | 359 (56.6) | **1200 (47.9)** |
| **Formula milk used in last 24-hours** | **523 (12.1)** | 69 (11.5) | 109 (18.2) | 106 (17.7) | **284 (15.8)** | 143 (22.9) | 53 (8.2) | 22 (3.7) | 21 (3.3) | **239 (9.5)** |
| **Formula milk used in last 7-days** | **652 (15.1)** | 75 (12.5) | 133 (22.2) | 119 (19.8) | **327 (18.2)** | 183 (29.3) | 69 (10.6) | 50 (8.3) | 23 (3.6) | **325 (13.0)** |
| **Other milk use in last 24-hours** | **408 (9.5)** | 8 (1.3) | 116 (19.3) | 71 (11.8) | **195 (10.8)** | 32 (5.1) | 39 (6.0) | 54 (9.0) | 88 (13.9) | **213 (8.5)** |
| **Other milk use in last 7-days** | **537 (12.5)** | 11 (1.8) | 153 (25.5) | 83 (13.8) | **247 (13.7)** | 43 (6.9) | 51 (7.9) | 77 (12.8) | 119 (18.8) | **290 (11.6)** |
| **Water-based liquids in last 24 hrs** | **564 (13.1)** | 121 (20.2) | 32 (5.3) | 222 (37.0) | **375 (20.8)** | 13 (2.1) | 36 (5.6) | 49 (8.2) | 91 (14.4) | **189 (7.5)** |
| **Water-based liquids in last 7-days<sup>y</sup>** | **798 (18.5)** | 169 (28.2) | 52 (8.7) | 272 (45.3) | **493 (27.4)** | 19 (3.0) | 62 (9.6) | 79 (13.2) | 145 (22.9) | **305 (12.2)** |
| **Solid/soft food in last 24-hours<sup>φ</sup>** | **552 (12.8)** | 175 (29.2) | 73 (12.2) | 110 (18.3) | **358 (19.9)** | 29 (4.6) | 45 (6.9) | 52 (8.7) | 68 (10.7) | **194 (7.7)** |
| **Solid/soft food in last 7-days<sup>†</sup>** | **814 (18.9)** | 191 (31.8) | 121 (20.2) | 138 (23.0) | **450 (25.0)** | 38 (6.1) | 56 (8.6) | 170 (28.4) | 100 (15.8) | **364 (14.5)** |
| **Prevalence of use of soft/solid food in the past 24-hours amongst all infants, by age groups, N (%)** |  |  |  |  |  |  |  |  |  |  |
| 0-30 days | **11 (1.7)** | 4 (4.0) | 0 | 4 (4.0) | **8 (2.7)** | 0 | 0 | 1 (1.0) | 2 (1.9) | **3 (0.8)** |
| 31-60 days | **18 (2.5)** | 7 (7.0) | 4 (4.0) | 3 (3.0) | **14 (4.7)** | 0 | 0 | 1 (1.0) | 3 (2.7) | **4 (1.0)** |
| 61-90 days | **41 (5.7)** | 18 (18.0) | 4 (4.0) | 5 (5.0) | **27 (9.0)** | 0 | 2 (1.9) | 8 (8.3) | 4 (3.9) | **14 (3.3)** |
| 91-120 days | **81 (11.1)** | 36 (36.0) | 6 (6.0) | 17 (17.0) | **59 (19.7)** | 1 (0.8) | 4 (4.0) | 9 (9.0) | 8 (7.6) | **22 (5.1)** |
| 121-150 days | **136 (18.8)** | 43 (43.0) | 20 (20.0) | 30 (30.0) | **93 (31.0)** | 5 (4.8) | 14 (12.2) | 14 (14.3) | 10 (9.6) | **43 (10.2)** |
| 151-180 days | **265 (35.1)** | 67 (67.0) | 39 (39.0) | 51 (51.0) | **157 (52.3)** | 23 (15.5) | 25 (24.3) | 19 (18.6) | 41 (39.8) | **108 (23.7)** |
| Not known | **1** | 0 | 0 | 0 | **0** | 0 | 1 | 0 | 0 | **1** |
| **Prevalence of use of soft/solid food in the past 7-days amongst all infants, by age groups, N (%)** |  |  |  |  |  |  |  |  |  |  |
| 0-30 days | **14 (2.1)** | 4 (4.0) | 1 (1.0) | 4 (4.0) | **9 (3.0)** | 0 | 0 | 3 (3.0) | 2 (1.9) | **5 (1.4)** |
| 31-60 days | **37 (5.1)** | 9 (9.0) | 5 (5.0) | 6 (6.0) | **20 (6.7)** | 0 | 0 | 14 (13.6) | 3 (2.7) | **17 (4.0)** |
| 61-90 days | **65 (9.0)** | 21 (21.0) | 9 (9.0) | 7 (7.0) | **37 (12.3)** | 0 | 2 (1.9) | 22 (22.9) | 4 (3.9) | **28 (6.6)** |
| 91-120 days | **132 (18.1)** | 41 (41.0) | 13 (13.0) | 26 (26.0) | **80 (26.7)** | 1 (0.8) | 5 (5.0) | 33 (33.0) | 13 (12.3) | **52 (12.1)** |
| 121-150 days | **202 (28.0)** | 47 (47.0) | 33 (33.0) | 36 (36.0) | **116 (38.7)** | 6 (5.7) | 19 (16.5) | 40 (40.8) | 21 (20.2) | **86 (20.4)** |
| 151-180 days | **364 (48.2)** | 69 (69.0) | 60 (60.0) | 59 (59.0) | **188 (62.7)** | 31 (21.0) | 30 (29.1) | 58 (56.9) | 57 (55.3) | **176 (38.6)** |
| Not known | 2 | 0 | 0 | 0 | 0 | 0 | 1 | 1 | 0 | 2 |
| Traditional medication used in last 24-hours<sup>†</sup> | 414 (9.6) | 76 (12.7) | 158 (26.3) | 109 (18.2) | 343 (19.1) | 1 (0.2) | 42 (6.5) | 8 (1.3) | 20 (3.2) | 71 (2.8) |
| Traditional medication used in last 7-days | 570 (13.2) | 90 (15.0) | 229 (38.2) | 135 (22.5) | 454 (25.2) | 2 (0.3) | 59 (9.1) | 27 (4.5) | 28 (4.4) | 116 (4.6) |
| Any prescribed medication used in last 24-hours<sup>‡</sup> | 1054 (24.5) | 171 (28.5) | 384 (64.0) | 165 (27.5) | 720 (40.0) | 13 (2.1) | 195 (30.1) | 35 (5.9) | 91 (14.4) | 334 (13.3) |
| Any prescribed medication used in last 7-days, N (%)● | 1396 (32.4) | 231 (38.5) | 451 (75.2) | 218 (36.4) | 900 (50.0) | 25 (4.0) | 258 (39.8) | 67 (11.2) | 146 (23.0) | 496 (19.8) |

\*Missing data Tanzania:1; †missing data: Nigeria: 1 Uganda: 1 ‡missing data: Nigeria:1 †: missing data Nigeria = 1; Tanzania=1 ‡missing Tanzania:2 †missing data n=2: Tanzania ‡ missing data: Tanzania n=4 ●missing data from Pakistan n=1

Across all sites, in the last 24-hours and 7-days, formula milk was consumed by 12.1% (95% CI 11.2-13.2%) and 15.1% (95% CI 14.1-16.2%) of infants respectively, with 1.5 times lower use in African (9.5%, 95% CI 8.4-10.8% and 13.0%, 95% CI 11.7-14.3% using 24-hour and 7-day recall, respectively) compared with Asian (15.8%, 95% CI 14.1-17.6% and 18.2%, 95% CI 16.4-20.0% at 24-hours and 7-days, respectively) sites, Table 3. However, since birth almost one-quarter of infants (23.9%, 95% CI 22.7-25.2%) had ever received formula milk, with higher use in Asia (31.6%, 95% CI 29.5-33.8%) than Africa (18.4%, 16.9-20.0%), Table 4, especially India (41.9%, 95% CI 37.9-46.0%) and Pakistan (39.0%, 95% CI 35.1-43.0%) compared with Bangladesh (14.0% (95% CI 11.3-17.0%)). In Africa, formula milk use was highest in Ethiopia (42.5%, 95% CI 38.6-46.5%), followed by Nigeria (16.2%, 95% CI 13.4- 19.2%), Tanzania (10.2%, 95% CI 7.9-12.9%), and Uganda (4.7%, 95% 3.2-6.7%). Most formula use started within the first month of life (65.0%), predominantly due to perceived insufficient breast milk (64.7%) and advice from healthcare providers (52.0%). Strikingly, only 46.9% of mothers using formula milk prepared it correctly, with substantial variation across sites from as low as 8.3% in Bangladesh to 61.6% in Pakistan and 77.0% in Ethiopia, Table 4. The details of other milk usage and traditional medicine usage (Tables 3 and 4), are described in the Supplementary material

**Table 4:**
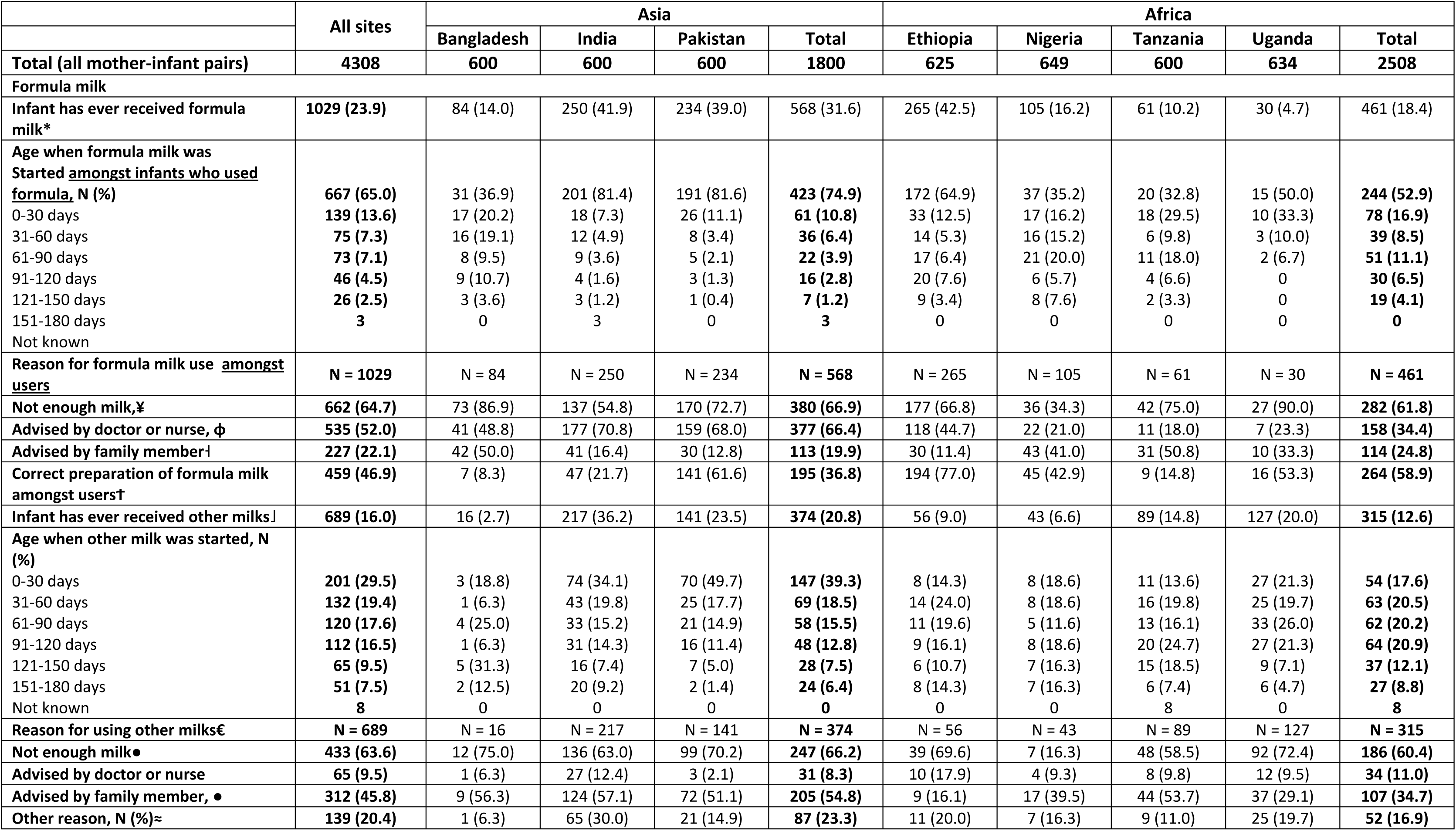
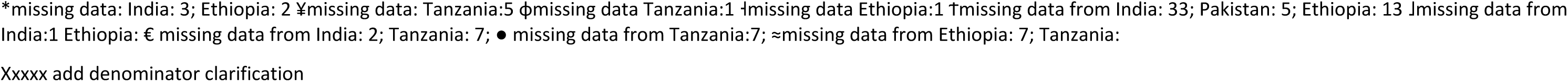
Use of formula milk and other milk Number (%)

|  | All sites | Asia |  |  |  | Africa |  |  |  |  |
| --- | --- | --- | --- | --- | --- | --- | --- | --- | --- | --- |
|  |  | Bangladesh | India | Pakistan | Total | Ethiopia | Nigeria | Tanzania | Uganda | Total |
| **Total (all mother-infant pairs)** | **4308** | **600** | **600** | **600** | **1800** | **625** | **649** | **600** | **634** | **2508** |
| **Formula milk** |  |  |  |  |  |  |  |  |  |  |
| **Infant has ever received formula milk\*** | **1029 (23.9)** | 84 (14.0) | 250 (41.9) | 234 (39.0) | 568 (31.6) | 265 (42.5) | 105 (16.2) | 61 (10.2) | 30 (4.7) | 461 (18.4) |
| **Age when formula milk was Started amongst infants who used formula, N (%)** | **667 (65.0)** | 31 (36.9) | 201 (81.4) | 191 (81.6) | **423 (74.9)** | 172 (64.9) | 37 (35.2) | 20 (32.8) | 15 (50.0) | **244 (52.9)** |
| 0-30 days | **139 (13.6)** | 17 (20.2) | 18 (7.3) | 26 (11.1) | **61 (10.8)** | 33 (12.5) | 17 (16.2) | 18 (29.5) | 10 (33.3) | **78 (16.9)** |
| 31-60 days | **75 (7.3)** | 16 (19.1) | 12 (4.9) | 8 (3.4) | **36 (6.4)** | 14 (5.3) | 16 (15.2) | 6 (9.8) | 3 (10.0) | **39 (8.5)** |
| 61-90 days | **73 (7.1)** | 8 (9.5) | 9 (3.6) | 5 (2.1) | **22 (3.9)** | 17 (6.4) | 21 (20.0) | 11 (18.0) | 2 (6.7) | **51 (11.1)** |
| 91-120 days | **46 (4.5)** | 9 (10.7) | 4 (1.6) | 3 (1.3) | **16 (2.8)** | 20 (7.6) | 6 (5.7) | 4 (6.6) | 0 | **30 (6.5)** |
| 121-150 days | **26 (2.5)** | 3 (3.6) | 3 (1.2) | 1 (0.4) | **7 (1.2)** | 9 (3.4) | 8 (7.6) | 2 (3.3) | 0 | **19 (4.1)** |
| 151-180 days | **3** | 0 | 3 | 0 | **3** | 0 | 0 | 0 | 0 | **0** |
| Not known |  |  |  |  |  |  |  |  |  |  |
| **Reason for formula milk use amongst users** | **N = 1029** | N = 84 | N = 250 | N = 234 | **N = 568** | N = 265 | N = 105 | N = 61 | N = 30 | **N = 461** |
| **Not enough milk,¥** | **662 (64.7)** | 73 (86.9) | 137 (54.8) | 170 (72.7) | **380 (66.9)** | 177 (66.8) | 36 (34.3) | 42 (75.0) | 27 (90.0) | **282 (61.8)** |
| **Advised by doctor or nurse, ϕ** | **535 (52.0)** | 41 (48.8) | 177 (70.8) | 159 (68.0) | **377 (66.4)** | 118 (44.7) | 22 (21.0) | 11 (18.0) | 7 (23.3) | **158 (34.4)** |
| **Advised by family member†** | **227 (22.1)** | 42 (50.0) | 41 (16.4) | 30 (12.8) | **113 (19.9)** | 30 (11.4) | 43 (41.0) | 31 (50.8) | 10 (33.3) | **114 (24.8)** |
| **Correct preparation of formula milk amongst users†** | **459 (46.9)** | 7 (8.3) | 47 (21.7) | 141 (61.6) | **195 (36.8)** | 194 (77.0) | 45 (42.9) | 9 (14.8) | 16 (53.3) | **264 (58.9)** |
| **Infant has ever received other milks‡** | **689 (16.0)** | 16 (2.7) | 217 (36.2) | 141 (23.5) | **374 (20.8)** | 56 (9.0) | 43 (6.6) | 89 (14.8) | 127 (20.0) | **315 (12.6)** |
| **Age when other milk was started, N (%)** |  |  |  |  |  |  |  |  |  |  |
| 0-30 days | **201 (29.5)** | 3 (18.8) | 74 (34.1) | 70 (49.7) | **147 (39.3)** | 8 (14.3) | 8 (18.6) | 11 (13.6) | 27 (21.3) | **54 (17.6)** |
| 31-60 days | **132 (19.4)** | 1 (6.3) | 43 (19.8) | 25 (17.7) | **69 (18.5)** | 14 (24.0) | 8 (18.6) | 16 (19.8) | 25 (19.7) | **63 (20.5)** |
| 61-90 days | **120 (17.6)** | 4 (25.0) | 33 (15.2) | 21 (14.9) | **58 (15.5)** | 11 (19.6) | 5 (11.6) | 13 (16.1) | 33 (26.0) | **62 (20.2)** |
| 91-120 days | **112 (16.5)** | 1 (6.3) | 31 (14.3) | 16 (11.4) | **48 (12.8)** | 9 (16.1) | 8 (18.6) | 20 (24.7) | 27 (21.3) | **64 (20.9)** |
| 121-150 days | **65 (9.5)** | 5 (31.3) | 16 (7.4) | 7 (5.0) | **28 (7.5)** | 6 (10.7) | 7 (16.3) | 15 (18.5) | 9 (7.1) | **37 (12.1)** |
| 151-180 days | **51 (7.5)** | 2 (12.5) | 20 (9.2) | 2 (1.4) | **24 (6.4)** | 8 (14.3) | 7 (16.3) | 6 (7.4) | 6 (4.7) | **27 (8.8)** |
| Not known | **8** | 0 | 0 | 0 | **0** | 0 | 0 | 8 | 0 | **8** |
| **Reason for using other milks€** | **N = 689** | N = 16 | N = 217 | N = 141 | **N = 374** | N = 56 | N = 43 | N = 89 | N = 127 | **N = 315** |
| **Not enough milk●** | **433 (63.6)** | 12 (75.0) | 136 (63.0) | 99 (70.2) | **247 (66.2)** | 39 (69.6) | 7 (16.3) | 48 (58.5) | 92 (72.4) | **186 (60.4)** |
| **Advised by doctor or nurse** | **65 (9.5)** | 1 (6.3) | 27 (12.4) | 3 (2.1) | **31 (8.3)** | 10 (17.9) | 4 (9.3) | 8 (9.8) | 12 (9.5) | **34 (11.0)** |
| **Advised by family member, ●** | **312 (45.8)** | 9 (56.3) | 124 (57.1) | 72 (51.1) | **205 (54.8)** | 9 (16.1) | 17 (39.5) | 44 (53.7) | 37 (29.1) | **107 (34.7)** |
| **Other reason, N (%)≈** | **139 (20.4)** | 1 (6.3) | 65 (30.0) | 21 (14.9) | **87 (23.3)** | 11 (20.0) | 7 (16.3) | 9 (11.0) | 25 (19.7) | **52 (16.9)** |

\*missing data: India: 3; Ethiopia: 2 ✕missing data: Tanzania:5 ϕmissing data Tanzania:1 †missing data Ethiopia:1 †missing data from India: 33; Pakistan: 5; Ethiopia: 13 †missing data from India:1 Ethiopia: € missing data from India: 2; Tanzania: 7; ● missing data from Tanzania:7; ≈missing data from Ethiopia: 7; Tanzania:
Xxxxx add denominator clarification

One in five mothers (20.8%, 95% CI 19.6-22.0%) reported at least one breastfeeding difficulty, Table 5 and Supplementary table S1, the commonest being perceived milk insufficiency (17.2%, 95% CI 16.1-18.4%) and reported by almost a quarter of women in Asia compared with less than one-eighth in Africa, Table 5. Other difficulties included painful or tender breasts (2.9%, 95% CI 2.4-3.4%), sore nipples (2.8%, 95% CI 2.4-3.4%), and swollen breasts (1.3%, 95% CI 1.0-1.7%). Pakistan recorded the highest prevalence of breastfeeding difficulties (29.5% 95% CI 25.9-33.4%), followed by India, and Bangladesh, Table 5.

**Table 5:** Breastfeeding difficulties experienced amongst all women in the previous 7-days across seven countries in Asia and Africa Number (%)

|  | All sites | Asia |  |  |  | Africa |  |  |  |  |
| --- | --- | --- | --- | --- | --- | --- | --- | --- | --- | --- |
|  |  | Bangladesh | India | Pakistan | Total | Ethiopia | Nigeria | Tanzania | Uganda | Total |
| **Total mother-infant pairs** | **4308** | **600** | **600** | **600** | **1800** | **625** | **649** | **600** | **634** | **2508** |
| **Mother reports breastfeeding difficulties, N (%)\*** | 889 (20.8) | 146 (24.3) | 163 (27.4) | 175 (29.5) | **484 (27.1)** | 32 (5.1) | 68 (10.5) | 98 (16.6) | 207 (32.9) | **405 (16.3)** |
| Not having enough milk, N (%)† | 739 (17.2) | 126 (21.0) | 147 (24.7) | 168 (28.3) | **441 (24.7)** | 26 (4.2) | 35 (5.4) | 62 (10.4) | 175 (27.7) | **298 (11.9)** |
| Sore nipples, N (%)‡ | **122 (2.8)** | 5 (0.8) | 16 (2.7) | 10 (1.7) | **31 (1.7)** | 6 (1.0) | 26 (4.0) | 45 (7.5) | 14 (2.2) | **91 (3.6)** |
| Swollen breast(s) , N (%)¥ | **5 (1.3)** | 2 (0.3) | 2 (0.3) | 5 (0.8) | **9 (0.5)** | 5 (0.8) | 6 (0.9) | 19 (3.2) | 16 (2.5) | **46 (1.8)** |
| Painful or tender breasts, N (%)~ | **124 (2.9)** | 18 (3.0) | 14 (2.4) | 12 (2.0) | **44 (2.5)** | 7 (1.1) | 16 (2.5) | 28 (4.7) | 29 (4.6) | **80 (3.2)** |

\*Missing data: India: 4; Pakistan: 7, Ethiopia: 2; Nigeria: 3; Tanzania: 9; Uganda:4; † missing data: India: 4 Pakistan 7; Ethiopia: 2; Nigeria: 2; Tanzania:4; Uganda: 1; ‡ missing data for India: 4, Pakistan: 7, Ethiopia:2, Nigeria:1, Tanzania: 3, Uganda:1; ¥ missing data for India:4, Pakistan:7, Ethiopia: 2, Nigeria:1, Tanzania: 1, Uganda:1; ~missing data on India: 4, Pakistan: 7, Ethiopia:2, Nigeria:2, Tanzania:1, Uganda:

## Discussion

Our most striking findings were that despite the high proportion of facility-based deliveries, and almost universal uptake of any breastfeeding, EIBF occurred in only half the study population, and was 1.5 times higher in African versus Asian settings; most interesting was the subtle paradox that in Asia, EIBF was highest in Bangladesh (48%) despite 44% of infants being born at home, while in sites with more than 90% in-facility deliveries (India, Ethiopia, and Tanzania), EIBF was 23%, 52% and 83%, respectively; furthermore, EBF prevalence was low (40% overall), with introduction of formula and other milks within the first 60-days of life, mainly due to perceived milk insufficiency or following advice from health workers; most concerning was that less than half of formula milk users prepared it safely. Surprisingly, very few mothers reported a physical breastfeeding difficulty such as sore nipples or swollen breasts. Using 7-day recall data, the WHA 2025 50% EBF target was achieved by Ethiopia, Nigeria and Uganda, but the Global Collective’s 60% EBF target was not achieved by any site.(13)

The low EIBF prevalence we measured is consistent with DHS data which measured 51.9% (95% CI 51.6–52.2) EIBF prevalence (2010–2018), with higher EIBF in Africa (55.8% (95% CI 55.3–56.3)) versus South East Asia / Western Pacific (47.4 (46.9–47.9)).(22) Our EIBF finding aligns with the 2023 Lancet Breastfeeding series which described EIBF in less than 50% of children, and prelacteal feeds in one-third, globally, 2010-2019.(23) The low EBF prevalence (40.2% (95% CI 38.7-41.7%)), we measured is lower than the 45.7% (95% CI (45.2–46.2)) global weighted 0-6 month EBF prevalence measured using DHS data, from 57 LMICs, 2010-2018, (22) and similar to the 38.7% (28.3–49.9%) measured in LMICs in 2018.(24) Contrary to DHS findings we describe higher EBF prevalence in African, versus Asian sites, (DHS: EBF 41.7 (41.0–42.4) in Africa compared with 55.2 (54.4–56.0) in South East Asia / Western Pacific regions) (22), possible because we included well-supported sites in Africa, but the very low EBF prevalence in Pakistan is similar across data sets, and is concerning. Other analyses projected an increase in overall, EBF prevalence in LMICs from 38.7% (28.3– 49.9%) in 2018 to 42.6% (25.6–60.5%) in 2025 – close to our measurement, and modelling estimated that 45.2% EBF (23.9–67.2%) by 2030; (24) However, all confidence intervals overlap in these estimates. DHS data show that the 44 LMICs with prior data, reported a 10.1% increase in EBF prevalence, from 36.5% in 2000-2009 to 46.6%, 2010 to 2018, but no confidence intervals are provided. These data show that over 4-5 years, despite the 2016 Lancet Breastfeeding series that explains the critical role of breastfeeding for children and society, (25) and the 2023 Lancet breastfeeding series that exposed the political economy of breastfeeding and the predatory nature of the baby milk industry (26), we have not managed to significantly improve breastfeeding practices within health systems.

Despite this bleak outlook, it is reported that during1990-2019 the years of life lost (YLL) due to inadequate breastfeeding among children under five, decreased by 70% and the disparity between high- and low-income regions narrowed by 48% for diarrheal diseases and 13% for lower respiratory infections, possible due to improved breastfeeding practices.(27) These data show that impact may be possible, despite slow progress. Furthermore, secondary analyses of cross-sectional data from India found that EBF prevalence was higher when mothers were exposed to media, had normal BMI, and visited health centres > 4 times during pregnancy.(28)

We know that breastfeeding practices are sub-optimal due to maternal employment, fears of milk sufficiency, family, societal and structural barriers (including gender and power imbalances), and predatory marketing of breastmilk substitutes (23, 25, 26, 29) Furthermore, in Africa and Asia socio-cultural factors include beliefs that colostrum is dirty; the need to cleans the baby’s gut, quench thirst and flush the bladder; the role of significant others (grandmothers and husbands), and characteristics such as young or older maternal age, low level of education, being unmarried, low income, being employed or unemployed (depending on context), late breastfeeding initiation, female infant and being a primigravida.(28, 30) We found that EBF is displaced by fears of milk insufficiency, fuelled by access to breastmilk substitutes, which are often marketed using predatory techniques.(31, 32) One systematic review (2022) found that the sustainability of breastfeeding interventions was facilitated by having a strong facilitator and hindered by having low community health worker involvement.(3) For more than 20 years we have known that breastfeeding practices can be improved at a population level through individualised peer counselling and support, in addition to multilevel and multicomponent context-specific interventions.(23, 33) Breastfeeding counselling and support requires collective societal approaches that take gender inequities into consideration, and involve the family or community (4, 23), and enhances female empowerment, regardless of years of education.(34) WHO recommends strong investments in breastfeeding counselling and support (15), at the lowest level of the health system, and at home, (33, 35) free from commercial influences that promote breastmilk substitutes.(32) Political commitment, advocacy, financial investments and monitoring of the International Code of marketing of breastmilk substitutes, followed by action when contraventions are identified, are paramount.(32)

### Strengths and Limitations

Our survey’s major strengths include its inclusion of Asian and African countries and six infant age groups within the critical first six months of life. The large sample size enhances the reliability of findings allowing for robust statistical precision. Furthermore data collection used standardised instruments and training and field staff supervision. These enhance the generalizability and validity of the study’s conclusions across different cultural and regional contexts. Limitations include the cross-sectional design, potential recall and social desirability bias.

### Implications and conclusion

Our findings point to the critical need for context-specific breastfeeding counselling and support strategies for early and exclusive breastfeeding. These should build mothers’ and families’ confidence, strengthen breastfeeding support within the heath system and curb unnecessary formula milk marketing that exploits the fears, and concerns of parents and families.

## Data Availability

The data will be made available upon request to WHO, accompanied by a concept note explaining the reason for the request, and analysis plan. All site PIs will need to grant permission before the pooled data is shared.

## Acknowledgements

We acknowledge the mothers and infants who participated in this study, and the authors listed below:

|  | **First Name Middle initial(s) (optional) Last Name.** | **<u>Institution and Department</u>** | 1. Wrote protocol<br>2. Contributed to protocol development and finalisation<br>3. Contributed to SOPs / developed local SOPs<br>4. Data management (global or site level)<br>5. Oversaw data collection<br>6. Conducted Statistical analysis<br>7. Contributed to data interpretation<br>8. Drafted the manuscript<br>9. Contributed to the manuscript for important intellectual content<br>10. Approved final version of manuscript |
| --- | --- | --- | --- |
| Bangladesh | Abdullah H. Baqui | Department of International Health, Johns Hopkins Bloomberg School of Public Health | 2, 3, 5, 7, 8, 9, 10 |
|  | Rukhsana Haider | Projahnmo Research Foundation | 2, 3, 5, 7, 8, 9, 10 |
|  | Salahuddin Ahmed | Projahnmo Research Foundation | 2, 3, 5, 7, 8, 9, 10 |
|  | Ahad Mahmud Khan | Projahnmo Research Foundation | 2, 3, 4, 5, 7, 9, 10 |
|  | Kayur Mehta | Department of International Health, Johns Hopkins Bloomberg School of Public Health | 2, 3, 4, 7, 9, 10 |
|  | Md Shafiqul Islam | Projahnmo Research Foundation | 4,5,9,10 |
|  | Nabidul Haque Chowdhury | Projahnmo Research Foundation | 4, 5, 7, 8, 10 |
| Ethiopia | Yemane Berhane, | Department of Public Health, Addis Continental Institute of Public Health | 2, 3, 5, 7, 8, 9, 10 |
|  | Semira Abdelmenan, | Department of Public Health, Addis Continental Institute of Public Health | 2, 3, 4,5, 6, 7, 8, 9, 10 |
|  | Dagmawit Tewahido, | Department of Public Health, Addis Continental Institute of Public Health | 2,3,7,8,9,10 |
|  | Hanna Berhane, | Department of Public Health, Addis Continental Institute of Public Health | 2,3,7,8,9,10 |
|  | Wafaie Fawzi, | Departments of Nutrition; Epidemiology ; and Global Health and Population, Harvard T.H. Chan School of Public Health, Boston, Massachusetts, USA | 2,7,8,9,10 |
|  | Uttara Partap, | Department of Global Health and Population, Harvard T.H. Chan School of Public Health, Boston, Massachusetts, USA | 2,7,8,9,10 |
|  | Dongqing Wang | Department of Global and Community Health, College of Public Health, George Mason University, Fairfax, Virginia, USA | 2,7,8,9,10 |
| India | Sowmya C Karantha, | Society for Applied Studies | 2,3,8,9,10 |
|  | Sitanshi Sharma, | Society for Applied Studies | 2,3,8,9,10 |
|  | Kiran Bhatia, | Society for Applied Studies | 4,5,6 |
|  | Ranadip Chowdhury, | Society for Applied Studies | 2,3,4,5,6,7,8,9,10 |
|  | Harish Chellani, | Society for Applied Studies | 9,10 |
|  | Sunita Taneja, | Society for Applied Studies | 2,3,4,5,9,10 |
|  | Nita Bhandari | Society for Applied Studies | 2,3,9,10 |
| Nigeria | Adejuyigbe Ebunoluwa A, | University of Medical Sciences, Ondo City, Nigeria | 2, 3, 4, 5, 7, 8, 9, 10 |
|  | Ojo Temitope, | Department of Community Medicine, Obafemi Awolowo University, Ile-Ife, Nigeria. | 2, 3, 4, 5, 7, 8, 9, 10 |
|  | Titilayo Ayotunde, | Department of Demography and Statistics, Obafemi Awolowo University, Ile-Ife, Nigeria. | 3, 4, 5, 7, 8, 9, 10 |
|  | Anyabolu Henry, | Department of Paediatrics and Child health, Obafemi Awolowo University, Ile-Ife, Nigeria | 2, 3, 7, 8, 9, 10 |
|  | Ogundele Tolulope | Wesley Guild Hospital, Ilesa, , Nigeria | 2,3,,5,7 |
|  | Mohammed Amina | Child Health Division, Federal Ministry of Health., Nigeria. | 3, 5, 7, 9 |
|  | Awowole Ibrahim | Department of Obstetrics and gynaecology, Obafemi Awolowo University, Ile=Ife | 3, 5, 7, 9 |
| Pakistan | Dr. Fyezah Jehan, | Aga Khan University Department of Paediatrics and Child Health | 2, 3, 5, 7, 8, 9, 10 |
|  | Dr. Imran Nisar, | Aga Khan University<br>Department of Paediatrics<br>and Child Health | 2, 3, 5, 7, 8, 9, 10 |
|  | Usma Mehmood, | Aga Khan University<br>Department of Paediatrics<br>and Child Health | 2, 3, 4, 5, 7, 9, 10 |
|  | Aysha Arif | Aga Khan University<br>Department of Paediatrics<br>and Child Health | 2, 3, 4, 5, 7, 9, 10 |
|  | Junaid Mehmood | Aga Khan University<br>Department of Paediatrics<br>and Child Health | 4,6,7 |
|  | Amina Barkat | Aga Khan University<br>Department of Paediatrics<br>and Child Health | 3,5,8 |
|  | Salman Osmani | Aga Khan University<br>Department of Paediatrics<br>and Child Health | 4,6,7 |
| Tanzania | Dr Usha Dhingra | Center for Public Health<br>Kinetics, Delhi, India | 2,3,4,5,6,7,8,9,10 |
|  | Arup Dutta | Center for Public Health<br>Kinetics, Delhi, India and<br>Swami Vivekanand<br>Subharti University, Meerut,<br>India | 3,4,5 |
|  | Fahad Aftab | Center for Public Health<br>Kinetics, Delhi, India | 4,5 |
|  | Siti Makame Ali | Center for Public Health<br>Kinetics, Delhi, India | 4,5 |
|  | Beshara Ali Alawi | Center for Public Health<br>Kinetics, Delhi, India | 4,5 |
|  | Dr. Mayassa Salum<br>Ally | Zanzibar Health Research<br>Institute, Zanzibar,<br>Tanzania | 3,5,10 |
|  | Dr Sunil Sazawal | Swami Vivekanand<br>Subharti University, Meerut,<br>India; Center for Public<br>Health Kinetics, Delhi,<br>India | 2,3,5,7,9,10 |
| Uganda | Grace Ndeezi, | Department of Paediatrics<br>and Child Health, College<br>of Health Sciences,<br>Makerere University | 2,3,4,5,6,7,8,9,10 |
|  | Dan Kajungu, | Makerere University<br>Centre for Health &<br>Population Research;<br>Department of Global<br>Health, Stellenbosch<br>University, South Africa | 2,3,4,5,6,7,8,9,10 |
|  | Nicolette Nabukeera<br>, | Department of Paediatrics<br>and Child Health, College<br>of Health Sciences,<br>Makerere University | 2,3,4,5,6,7,8,9,10 |
|  | Amanda Murungi, | Mulago National Referral<br>Hospital | 2,3,4,5,8,9,10 |
|  | Ezekiel Mupere, | Department of Paediatrics<br>and Child Health, College<br>of Health Sciences,<br>Makerere University | 2,3,4,5,7,9,10 |
|  | Philipa Musoke |  | 2,3,4,5,7,9,10 |
|  | Yehu Taremwa | Makerere University<br>Centre for Health &<br>Population Research | 4, 6, 7, 8, 9, 10 |
| WHO Sponsor | Karen Edmond | Dept of Sexual,<br>Reproductive, Maternal,<br>Child, Adolescent and<br>Older Persons Health<br>(LHR) | 1, 2, 3, 4, 5, 7, 8, 9, 10 |
|  | Nigel Rollins |  | 1, 2, 3, 4, 5, 7, 8, 9, 10 |
|  | Ameena Goga |  | 1, 2, 3, 4, 5, 7, 8, 9, 10 |
| Epidemiologist | Lisa Hurt |  | 4, 6, 7, 8, 9, 10 |

Furthermore, we would like to thank the GATES Foundation who funded this work as part of INV-051706.

The authors would like to thank the following individuals who contributed to the success of the survey in each site:

Furthermore, we would like to thank the GATES Foundation who funded this work as part of INV-051706. The authors would like to thank the following individuals who contributed to the success of the survey in each site:
|  | **First Name Middle initial(s) (optional) Last Name.** | **Institution** | **Reason for acknowledgement** |
| --- | --- | --- | --- |
| Bangladesh | ASMD Ashraful Islam | Projahnmo Research Foundation | Supervise field implementation |
|  | Monzur Rahman Bhuiyan | Projahnmo Research Foundation | Supervise field implementation |
|  | Md. Shahidullah | Projahnmo Research Foundation | Supervise field implementation |
| Ethiopia | Yordanos Fekadu | Addis Continental Institute of Public Health | Supervise field implementation |
|  | Elsabet Wujira | Addis Continental Institute of Public Health | Supervise field implementation |
|  | Roman Getachew | Addis Continental Institute of Public Health | Supervise field implementation |
|  | Sitota Tsegaye | Addis Continental Institute of Public Health | Study inception and Supervise field implementation |
| India | Kriti Jain | Society for Applied Studies | Supervise field implementation |
|  | Jasmine Kaur | Society for Applied Studies | Supervise field implementation |
|  | Brinda Dube | Society for Applied Studies | Supervise field implementation |
|  | Geeta Mehto | Society for Applied Studies | Supervise field implementation |
|  | Ramanjeet Kaur | Society for Applied Studies | Supervise field implementation |
|  | Neelam Kaur | Society for Applied Studies | Supervise field implementation |
| Nigeria | Onaolapo Oluwaseun | Center for Health Research and Development Studies O | Supervise field implementation and Data management |
|  | Oluwatosin Awoyode | Center for Health Research and Development studies, Ile-ife, Osun State , Nigeria | Supervise Data Collection and Data Management |
| Pakistan | Zahra Hoodbhoy | Aga Khan University | Provides technical and managerial leadership for the Vital Registry system, a collaborative platform generating and maintaining comprehensive line lists of households and mother–child dyads across study sites |
|  | Taranum Naz | Aga Khan University | Supervise field implementation and monitoring |
|  | Ambreen Nadir Hussain | Aga Khan University | Supervise field implementation and monitoring |
| Tanzania | Maryam Fihri Said | Center for Public Health Kinetics, Delhi, India |  |
|  | Mwinyi Issa Msellem | Zanzibar Public Health Laboratory, Zanzibar, Tanzania |  |
| Uganda | Dr. Getrude Nakitende |  | Coordination and supervision of field implementation |
|  | Collins Gyezaho |  | Data management support |
|  | James Tumwine |  | Contributed to Protocol development |
|  | Sarah Kiguli |  | Contributed to Protocol development and SOPs |
|  | Jesca Nsungwa |  | Contributed to Protocol development |
|  | Peter Waiswa |  | Contributed to Protocol development and SOPs |

## References

1. United National Inter-agency group for child mortality estimations. Level and Trends in Child Mortality. New York; 2024. Available from https://data.unicef.org/resources/levels-and-trends-in-child-mortality-2024/. Accessed 1 November 2025.

2. World Health Organization, UNICEF. Nutrition for growth summit: Four smart breastfeeding commitments.2025. Available from https://www.globalbreastfeedingcollective.org/reports/2025-nutrition-growth-summit. Accessed 1 November 2025

3. Engelhart A, Mason S, Nwaozuru U, Obiezu-Umeh C, Carter V, Shato T, et al. Sustainability of breastfeeding interventions to reduce child mortality rates in low, middle-income countries: A systematic review of randomized controlled trials. Front Health Serv 2022;2:doi: 10.3389/frhs.2022.88939.

4. CJ. F. The immunological components of human milk and their effect on immune development in infants. . J Nutr 2005;135:1–4. 10.1093/jn/135.1.1.

5. Richard S, McCormick B, Seidman J, Rasmussen Z, Kosek M, Rogawski E, et al. Relationships among common illness symptoms and the protective effect of breastfeeding in early childhood in MAL-ED: an eight-country cohort study. . Am J Trop Med Hyg. 2018;98(904):10.4269/ajtmh.17-0457.

6. Victora C, Bahl R, Barros A, França G, Horton S, Krasevec J, et al. Breastfeeding in the 21st century: epidemiology, mechanisms, and lifelong effect. The Lancet. 2016;387(10017):475–90.

7. Rollins NC BN, Hajeebhoy N, Horton S, Lutter CK, Martines JC, Piwoz EG, Richter LM, Victora CG; . Lancet Breastfeeding Series Group. Why invest, and what it will take to improve breastfeeding practices? Lancet. 2016;387(10017):491–504. doi: 10.1016/S0140-6736(15)01044-2. PMID: 26869576.

8. Horta B, Victora C, for World Health Organization. Long-term effects of breastfeeding: a systematic review. World Health Organization. . 2013.

9. Dwiantini F, Pamungkasari E, Andriani R. Meta Analysis: Effect of Exclusive Breastfeeding on Child’s Development. . J Matern Child Health. 2023;09:47–61. 10.26911-/thejmch.2024.09.01.05.

10. Phukan D, Ranjan M, Dwivedi L, et al. Impact of timing of breastfeeding initiation on neonatal mortality in India. Journal: International Breastfeeding Journal. 2018;13(27): 10.1186/s13006-018-0162-0.

11. World Health Organization. Global health risks: mortality and burden of disease attributable to selected major risks.2009. Available from https://www.who.int/publications/i/item/9789241563871. Accesse 1 November 2025

12. Jain S, Ahsan S, Robb Z, Crowley B, Walters D. The cost of inaction: a global tool to inform nutrition policy and investment decisions on global nutrition targets. . Health Policy and Planning. 2024;39(8):819–30. 10.1093/heapol/czae056.

13. Global Breastfeeding Collective Breastfeeding Scorecard [Internet]. 2023 [cited 23 March 2025]. Available from: https://www.unicef.org/media/150586/file/Global%20breastfeeding%20scorecard%202023.pdf

14. World Health Organization and the United Nations Children’s Fund (UNICEF), 2021. Indicators for assessing infant and young child feeding practices: definitions and measurement methods. Geneva: Licence: CC BYNC-SA 3.0 IGO; https://creativecommons.org/licenses/by-nc-sa/3.0/igo.

15. World Health Organization. Guideline: counselling of women to improve breastfeeding practices. 2018. . Licence: CC BY-NC-SA 3.0 IGO.

16. Hossain S, Mihrshahi S. Effect of exclusive breastfeeding and other infant and young child feeding practices on childhood morbidity outcomes: associations for infants 0–6 months in 5 South Asian countries using Demographic and Health Survey data. . Int Breastfeed J 2024;19(35):10.1186/s13006-024-00644-x.

17. World Health Organization, UNICEF. Global Nutrition Targets 2025 Breastfeeding Policy Brief. 2014. Available from https://iris.who.int/server/api/core/bitstreams/7312ce84-0e54-46a5-8393-1fd241c318ed/content. Accessed 1 November 2025.

18. Infant and young child feeding. [Internet]. 2021 [cited 23 March 2025]. Available from: https://data.unicef.org/topic/nutrition/infant-and-young-child-feeding/.

19. Patel A, Badhoniya NK, S., Senarath U., Agho K, Dibley M. Infant and young child feeding indicators and determinants of poor feeding practices in India: A national cross-sectional survey. BMC Pediatrics. 2015;10(106):10.1186/471-2431-10-106.

20. Benedict R, Craig H, Torlesse H. Staying engaged: Working mothers and breastfeeding practices in South Asia. . Maternal & Child Nutrition. 2018;14: e12612. 10.1111/mcn.

21. World Health Organization. Marketing of breast-milk substitutes: National implementation of the international code. 2020. Available from https://www.who.int/publications/i/item/9789240094482 Accessed 1 November 205.

22. Zong Xn, Wu H, Zhao M, Magnussen CG, Xi B. Global prevalence of WHO infant feeding practices in 57 LMICs in 2010-2018 and time trends since 2000 for 44 LMICs. eClinicalMedicine. 2021;37.

23. Pérez-Escamilla R, Tomori C, Hernández-Cordero S, Baker P, Barros AJD, Bégin F, et al. Breastfeeding: crucially important, but increasingly challenged in a market-driven world. The Lancet. 2023;401(10375):472–85.

24. Bhattacharjee NV, Schaeffer LE, Hay SI, Lu D, Schipp MF, Lazzar-Atwood A, et al. Mapping inequalities in exclusive breastfeeding in low- and middle-income countries, 2000–2018. Nature Human Behaviour. 2021;5(8):1027–45.

25. 2016 Lancet Breastfeeding series [Internet]. 2016 [cited 4 September 2025]. Available from: https://www.thelancet.com/series-do/breastfeeding.

26. The Lancet Breastfeeding series 2023 [Internet]. 2023 [cited 4 September 2025]. Available from: https://www.thelancet.com/series-do/breastfeeding-2023.

27. Miatton A, Buja A, Cozzolino C, Mafrici SF, Zamagni G, Monasta L, et al. The global burden of suboptimal breastfeeding from 1990 to 2019: results and insights of the Global Burden of Disease. Eur J Pediatr. 2025;184(7):448.

28. Aggarwal R, Garg P, Verma M, Bindal P, Aditi A, Kaur I, et al. Decadal trends in the exclusive breastfeeding practices among working Indian mothers: a multi-level analysis. Int Breastfeed J. 2024;19(1):83.

29. Balogun OO, Dagvadorj A, Anigo KM, Ota E, Sasaki S. Factors influencing breastfeeding exclusivity during the first 6 months of life in developing countries: a quantitative and qualitative systematic review. Matern Child Nutr. 2015;11(4):433–51.

30. Amzat J, Aminu K, Matankari B, Ismail A, Almu B, Kanmodi KK. Sociocultural context of exclusive breastfeeding in Africa: A narrative review. Health Sci Rep. 2024;7(5):e2115.

31. The L. Unveiling the predatory tactics of the formula milk industry. The Lancet. 2023;401(10375):409.

32. Rollins N, Piwoz E, Baker P, Kingston G, Mabaso KM, McCoy D, et al. Marketing of commercial milk formula: a system to capture parents, communities, science, and policy. The Lancet. 2023;401(10375):486–502.

33. Haider R, Ashworth A, Kabir I, Huttly SRA. Effect of community-based peer counsellors on exclusive breastfeeding practices in Dhaka, Bangladesh: a randomised controlled trial. The Lancet. 2000;356(9242):1643–7.

34. Dehghani M, Kazemi A, Heidari Z, Mohammadi F. The relationship between women’s breastfeeding empowerment and conformity to feminine norms. BMC Pregnancy and Childbirth. 2023;23(1):287.

35. Chapman DJ, Morel K, Anderson AK, Damio G, Pérez-Escamilla R. Breastfeeding peer counseling: from efficacy through scale-up. J Hum Lact. 2010;26(3):314–26.

